# ViraLite: An Ultracompact HIV Viral Load Self-Testing System with Internal Quality Control

**DOI:** 10.1101/2025.04.01.25325036

**Authors:** Anthony J. Politza, Tianyi Liu, Aneesh Kshirsagar, Ming Dong, Md. Ahasan Ahamed, Muhammad Asad Ullah Khalid, Roland Jones, Uttara Seshu, Kathryn Risher, Casey N. Pinto, Yusheng Zhu, Samir K Gupta, Weihua Guan

## Abstract

The availability of effective antiretroviral therapy has made HIV manageable, provided patients have consistent access to routine viral load (VL) testing. Nonetheless, access to frequent VL testing remains limited. There is a need for accessible, user-friendly testing systems that allow people living with HIV (PLHIV) to monitor their VL more frequently and empower self-management. Here, we developed ViraLite, a sample-to-answer, compact, accessible, and battery-powered system for HIV viral load monitoring. The system is built upon a probe-based RT-LAMP assay that allows for multiplexed detection and quantification. An internal quality control targeting the RNase P was incorporated to enhance the reliability of the results. A software-reconfigurable real-time sensing system empowered by machine learning and a smartphone-guided protocol was developed in tandem to analyze the multiplexed assay. We analyzed 45 clinically archived samples using ViraLite and benchmarked our results against qRT-PCR, which showed 21 positive and 23 negative samples. Using our process control, ViraLite first identified 17 inconclusive samples that would otherwise be classified as negative. Then, ViraLite classified 14 out of 15 HIV-positive samples (93.3%) and 13 out of 13 HIV-negative samples (100%). The incorporation of RNase P as a process control increased the sensitivity of ViraLite from 66.66% to 93.33%, while maintaining a high specificity (100%). To assess the acceptance of ViraLite among PLHIV, we recruited 480 participants from online and three clinical sites to complete a survey. Over 86% of participants indicated ViraLite had benefits in convenience and privacy, on the other hand 61% of participants indicated concerns with test accuracy. The integration of compact hardware, a reliable assay, and smartphone guidance provides an accurate, easy to use system for PLHIV to self-manage their viral load and update their prescriptions frequently.

## Introduction

Advances in antiretroviral therapy (ART) have reduced HIV mortality rates and lowered transmission rates(Eisinger et al., 2019; Liu et al., 2025b). Highly active ART (HAART), a combination ART uses several individual drugs to suppress viral replication and has been widely adopted as the clinical standard(Amor et al., 2006; Davey et al., 1999; J Buzón et al., 2010). When viral suppression is maintained below the detectable level, the risk of HIV transmission has been shown to be significantly decreased(Tanser et al., 2013). Thanks to these advances, 30 million people living with HIV (PLHIV) received treatment in 2023(Chamie et al., 2021; Stevens et al., 2014a; Stevens and Marshall, 2010; Wang et al., 2016). Despite advancements in treatment, HIV presents a major public health concern due to the absence of a cure(UNAIDS, n.d.; WHO, 2023). As a result, the number of PLHIV is increasing as therapies continue to improve, creating demand for accurate and reliable testing to help PLHIV maintain viral suppression. Because of the high stigma related to HIV, VL monitoring must be highly specific, protecting the public from false positive results that lead to unnecessary psychological distress. On the other hand, VL monitoring must maintain excellent analytical sensitivity to distinguish low copy numbers from false negatives, otherwise there is an increased risk for viral transmission through patient lifestyles. Therefore, to protect public health, viral load (VL) tests must consistently exhibit high sensitivity and specificity across diverse settings and platforms(Deeks et al., 2015).

Effective viral load management hinges on chronic and precise monitoring of viral load (VL) to maintain suppression and reduce transmission(Drain et al., 2019a). Chronic monitoring is provided from regularly scheduled blood tests written every 3-6 months by the patient’s physician(Peter et al., 2017). These tests enable precise monitoring of viral load by utilizing nucleic acid tests (NAT). NATs are most compatible with VL monitoring because they offer improved sensitivity and specificity over the faster, cheaper and more convenient alternatives, such as antigen and antibody tests(Niemz et al., 2011). NATs consist of three stages: 1. collection, 2. preparation, and 3. Analysis (Li et al., 2021). For this application, HIV VL monitoring, sample collection starts with a 3-5 mL blood sample, a requirement to produce the necessary amount of plasma (1-3 mL) for the next step. Viral RNA extraction can be achieved using solid-phase or liquid-phase extractions(Lee et al., 2023; Liu et al., 2025a; Wierucka and Biziuk, 2014), both with similar performance, which yield isolated viral RNA in a stabilizing buffer. Last, the number of viral RNA copies is analyzed by quantitative polymerase chain reaction (qPCR) (Piatak et al., 1993), the gold standard molecular test for quantifying nucleic acids due to its reliability and robustness(M. A. Ahamed et al., 2025; Ahamed and Guan, 2025; Oliveira et al., 2021). While NATs have provided very accurate and sensitive tests for HIV VL management, the turnaround time from sample to answer can extend from several days to weeks, severely delaying prescription updates and requiring numerous visits (Chamie et al., 2021; Drain et al., 2019a; Jangam et al., 2013; Pai et al., 2012; Schito et al., 2012; Shafiee et al., 2015; Stevens et al., 2014b; Stevens and Marshall, 2010).

To offer more convenient testing with shorter turnaround times, point of care devices aim to bring NAT into urgent care clinics and infectious disease centers(Dorward et al., 2018). This is accomplished by integrating all three stages of NAT into single, automated devices (Md. A. Ahamed et al., 2025; Sloma et al., 2009). Several POC devices for HIV, such as the Roche Cobas Ampliprep assay and C6000 analyzer, have been developed and incorporated into clinical workflows. Similar assays and devices from Cepheid and Abbott provide comparable performance and throughput for HIV clinics (Mariani et al., 2020). These devices significantly reduce the turnaround times of traditional lab-based NAT. However, the convenience of these devices is mainly witnessed in the laboratory, providing higher throughput with less hands-on time. The sample volumes required pose a significant challenge to patient convenience and comfort, as a venous blood draw is still necessary. Therefore, these devices are not well-suited to improve patient convenience in healthcare centers.

To improve the convenience of HIV VL monitoring, several studies have focused on developing devices for at-home testing by utilizing reduced sample volumes and miniaturized electronics. Finger prick blood has been validated as a testing sample for HIV, but the small volumes involved (25-100 µl) require very high LODs and robust sample preparation. Successful at-home testing devices offer semi-automated workflows from finger prick blood using magnetic beads and PCR detection. However, these devices are not ideal for accessible testing due to the requirements of PCR and reliance on lab infrastructure. Loop mediated isothermal amplification (LAMP) (Notomi et al., 2000) offers an alternative to PCR, offering comparable LODs and reaction-times(Tomita et al., 2008). Some devices have successfully implemented LAMP assays toward at-home or portable testing for a variety of infectious diseases and environmental monitoring. Our previous works (Choi et al., 2018, 2016; Liu et al., 2023, 2022) demonstrated the speed and LOD of LAMP for HIV VL monitoring but identified the need for specific readout methods of LAMP to resolve the outstanding issues such as false positives and lack of multiplexing to rival PCR (Dong et al., 2024; Rolando et al., 2020). Multiplexing is needed for at-home tests using LAMP to enable internal quality control monitoring to mimic conventional PCR assays that implement multiplex controls to consistently demonstrate high sensitivity and specificity(Hoorfar et al., 2004; Rosenstraus et al., 1998), a requirement for VL monitoring to safeguard public health (Bourlet et al., 2020). There is an unmet need for an HIV multiplexed RT-LAMP assay that uses internal quality controls for consistently high sensitivity and specificity toward at-home VL monitoring for people living with HIV (PLHIV).

Here in this work, we present ViraLite, an ultracompact HIV VL system utilizing multiplexed RT-LAMP to introduce internal quality controls for personal testing. Our device provides an easily accessible, user-guided test by implementing multiplex LAMP, semi-automated electronics, and smartphone assistance. The system is built upon a probe-based, multiplexable reverse-transcriptase-loop-mediated isothermal amplification (RT-LAMP) assay which enables reliable processing control and VL quantification. A versatile, and scalable sensing system empowered by machine learning (ML) was also developed alongside the multiplex assay. From 50 µl of plasma, the smartphone-guided system can detect HIV and RNase P (a process control) in a multiplexed reaction. We analyzed 45 clinically archived samples using ViraLite and benchmarked our results against qRT-PCR, which showed 21 positive and 23 negative samples. With our process control, we identified 17 inconclusive samples that would otherwise be classified as negative. Then, ViraLite classified 14 out of 15 HIV-positive samples (93.3%) and 13 out of 13 HIV-negative samples (100%), demonstrating the need for process control incorporation with RT-LAMP assays. Using this approach improved the sensitivity of ViraLite from 66.66% to 93.33%, while maintaining a high specificity (100%). To assess the usability and acceptance of ViraLite among PLHIV, we recruited 480 participants from online and three clinical sites to complete a survey. ViraLite showed strengths in Convenience and Privacy with over 86% of participants indicating benefits in these areas. On the other hand, participants identified test Accuracy (61%) as the largest concern to using our system. By integrating compact hardware, a reliable assay, and smartphone guidance, ViraLite offers an accurate and user-friendly system for PLHIV to self-manage their viral load and frequently update their prescriptions.

## Results

### ViraLite Workflow

To enable at home use, we designed and validated a portable sample-to-answer platform for nucleic acid testing (**Figure *1*a**). We previously validated the use of plasma separation membranes(Liu et al., 2023) to collect samples and light-weight centrifuges for portable RNA extraction(Politza et al., 2024), therefore enabling us to develop a fully portable system. A smartphone app will guide the user through each stage: Sample Collection, RNA Extraction, and Amplification (See **Supplementary Video S1**). Last, the extracted sample will be detected using our new multiplex RT-LAMP assay. This assay uses custom designed probes to enhance the specificity of each assay for use in multiplex format (**Figure *1*b-c**). ViraLite maintains a minimal packed footprint (**Figure *1*d**), and introduces our next generation of fluorescence analyzer, showcasing a machine learning pipeline that uses four multi-spectral color chips to empower one-pot multiplexing (**Figure *1*e**). Amplification is conducted on ViraLite to provide real-time updates to the user through the Bluetooth connected app.

**Figure 1.**
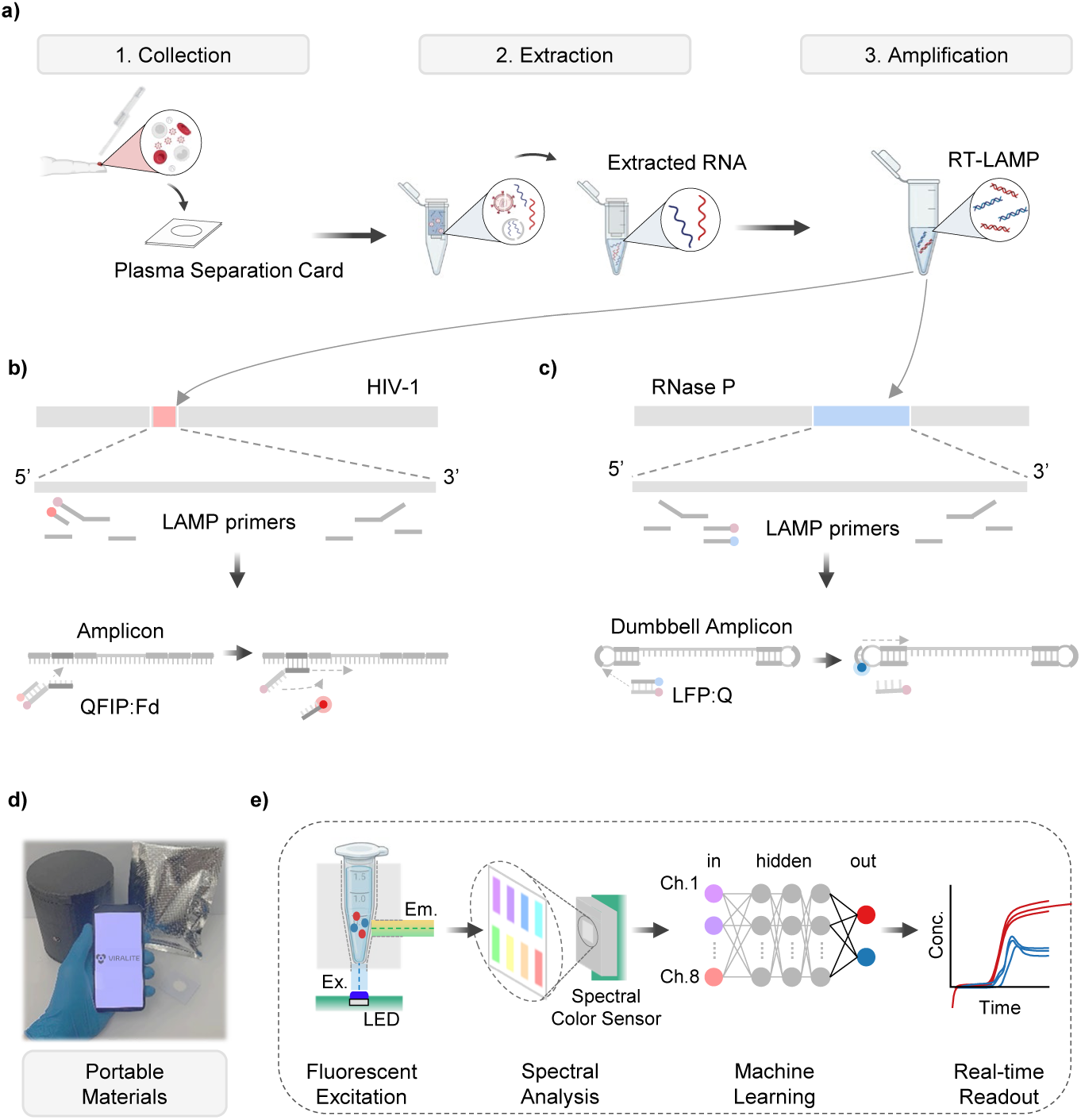
Demonstration of portable VL monitoring for self-management using the ViraLite protocols. a) Samples are first gathered using a plasma separation card that filters finger-prick blood and traps plasma into an absorption layer. Next, that layer is transferred to lysis buffer and RNA extraction is conducted using our lab developed portable centrifuge to process QIAamp viral RNA extractions. Last, the isolated RNA is added to our multiplex RT-LAMP assay for real-time analysis using our portable analyzer. b) The HIV-1 components of the multiplex assay target the reverse-transcriptase sub-region of the viral genome. A designed probe, using the FIP primer attached with a quencher, incorporates into the reaction as amplicons are generated and creates a change in fluorescence as a complementary fluorophore is released into the bulk assay. c) The Rnase P components of the multiplex assay targets the mRNA strand ‘RPP subunit p20’. Another designed probe, using the LF primer attached with a fluorophore, incorporates into the reaction as dumbbell amplicons are generated and creates a change in fluorescence as a complementary quencher is released into the bulk assay. d) The entire test can be packaged into a vacuum sealed pouch; extractions are verified for long-term storage without cold-chain dependency. The RT-LAMP assay still necessitates refrigeration. e) Detecting our multiplex assay is accomplished through a fine integration of hardware and software. The handheld device reads excited photons (480 nm Blue LED) from the bulk assay using a multi-spectra color sensor. The multi-spectra color sensor outputs 8-channels of usable RFU that can then be converted to dye/fluorophore concentration using a trained neural network (NN). Concentrations are then displayed to users via smartphone app or a provided summary tab.

### HIV RT-LAMP Assay with Internal Quality Control

To create an internal quality-controlled HIV assay, we designed two probe-based LAMP assays (**Supplementary Information**). We explored both assays in single-plex format before migrating into a one-pot multiplex assay (**Supplementary Figure S1**). The HIV RT-LAMP assay targets the Reverse-Transcriptase sub-region of the HIV genome and demonstrated a LOD of 21 copies (95% CI) (**Supplementary Figure S1a**). The RNase-P RT-LAMP assay targets the RNA strand ‘RPP subunit p20’ and demonstrated a LOD of 10 copies using a 95% confidence level (**Supplementary Figure S1b**). Both assays showed very strong correlation between benchtop and portable devices, suggesting suitable deployment into multiplex format (**Supplementary Figure S2a-b**).

To evaluate the cross-reactivity between our RT-LAMP assays and target samples, we analyzed the assay against three possible sample variations to measure specificity. We examined three scenarios: HIV and RNase P positive (to simulate viral rebound), only RNase P positive (to simulate a patient without viral rebound), and a No-target-control (NTC, to simulate an invalid test). Each sample was tested in triplicate using a one-pot multiplex assay. For each of these samples we found that only the appropriate target to channel relationship showed detection. For example, sample 1 showed both HIV and RNase P detection (**Figure 2a**). On the other hand, sample 2 showed only RNase P was detected. Sample 3 (a No Target Control) showed no detection for either target, demonstrating strong specificity and no primer interactions. These results were identical when tested using the ViraLite analyzer (**Supplementary Figure S3**). From these results we demonstrate that our multiplex assay is robust and capable of multiplex detection in clinically relevant scenarios.

**Figure 2.**
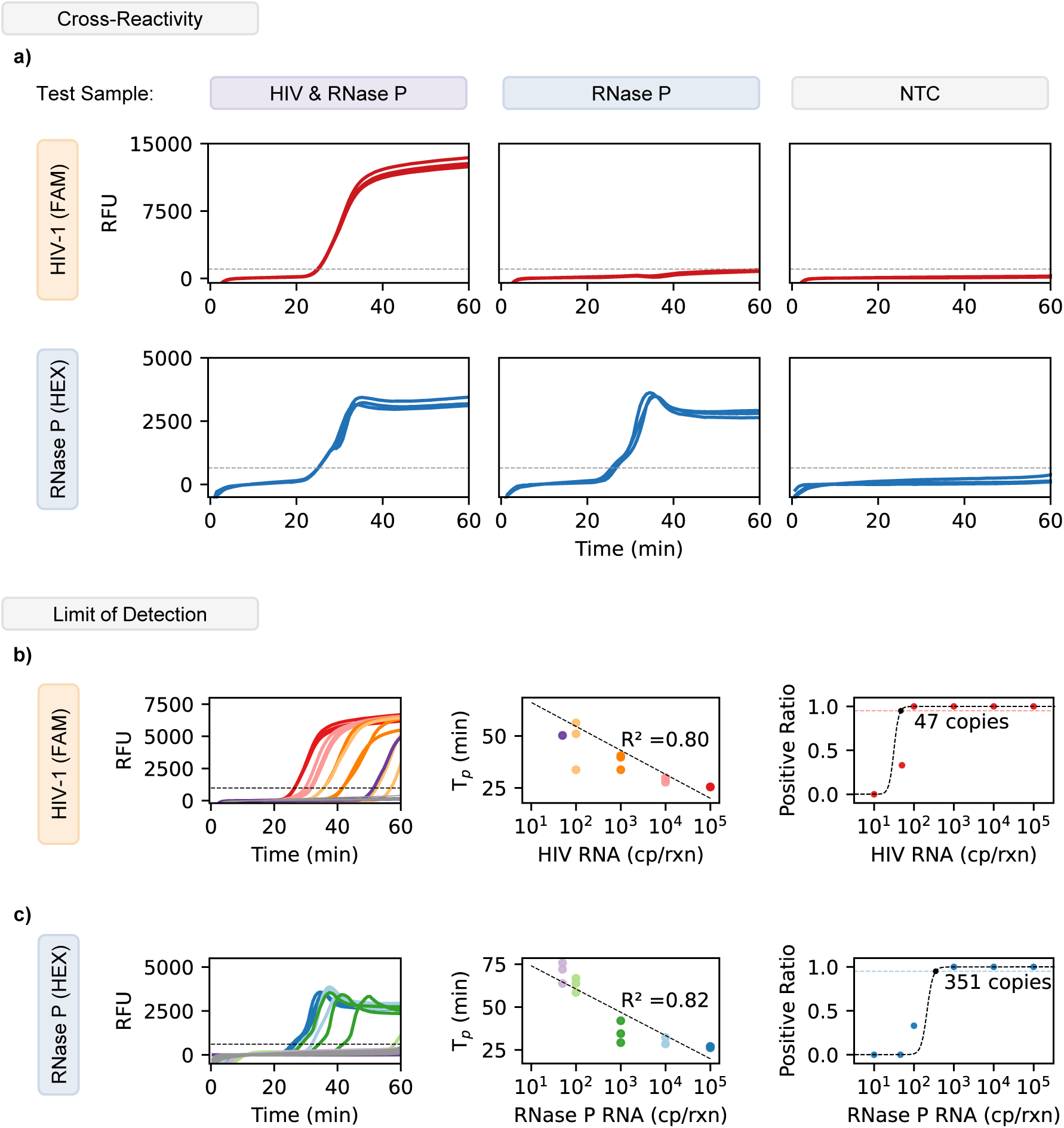
Validation of multiplex assay using cross-reactivity and limit of detection. a) The multiplex assay used FAM and HEX fluorophores to distinguish HIV-1 and RNase P, respectively. When both targets are present in the sample, there is exponential amplification for both FAM and HEX. For samples with only RNase P, there is only amplification detected in the HEX channel. No Target Controls (NTC) show no amplification for either FAM or HEX (HIV-1 or RNase P). b) Amplification curves using the multiplex assay to detect HIV-1 show positive detection from 10^5^ down to 47 cp/reaction. The multiplex assay shows a linear performance between HIV-1 RNA and Time to Positive (T_p_). The hit rate analysis for HIV-1 demonstrated a limit of detection (LOD) of 47 copies using the multiplex assay, this was established using a 95% positive ratio or higher. c) Amplification curves using the multiplex assay to detect RNase P show positive detection from 10^5^ down to 351 cp/reaction. The multiplex assay shows a linear performance between RNase P RNA and Time to Positive (T_p_). The hit rate analysis for RNase P demonstrated a limit of detection (LOD) of 351 copies using the multiplex assay, this was established using a 95% positive ratio or higher.

To investigate the limit of detection (LOD) of our multiplexed LAMP, we analyzed the multiplex assay with HIV-1 samples ranging from 10^5^ copies down to 10 copies per reaction. As shown in **Figure *2*b**, the multiplexed assay performed similarly to the single plex assay showing a reasonable R^2^ of 0.80 over the range of concentrations detected. The assay also showed a LOD of 47 copies. These results suggest that our assay has the potential for semi-quantitative abilities for HIV and can be used to process 100 µl samples which could contain concentrations as low as 1 cp/µl (1000 cp/mL). Next, we examined the RNase P with an identical range of RNA concentrations (10^5^ to 10 cp/rxn) (**Figure *2*c**). The multiplex assay showed similar performance to its single-plex counterpart for RNase P showing strong linearity (R^2^ = 0.82). Following a hit curve analysis, we found the LOD of the multiplex assay for RNase P was 351 copies. Despite a higher limit than anticipated, we still expect our multiplex assay to detect RNase P from plasma samples (∼12 cp/µl) (**Supplementary Figure S4**). Given a finger prick sample between 50-100 µl, we expect between 600-1200 copies, a value well above our multiplex LOD for RNase P. These results demonstrate our multiplex assay’s ability to be used for the quantitative detection of two targets simultaneously. Therefore, we can expect our assay to maintain high sensitivity and specificity when examining clinical samples using a portable device

### Validation of Multiplex Detection

To achieve a compact diagnostic device capable of multiplexed detection, we trained, tested, and integrated a neural network that converts 8-channel fluorescence into dye concentrations (FAM and HEX). Our previous study showed that a neural network worked best to clarify signal from overlapping emission spectra(Kshirsagar et al., 2024a) (**Supplementary Figure S5**), therefore enabling our device to be battery powered, lightweight, and easy to use while maintaining a highly accurate optical setup (**Figure *3*a)**(Choi et al., 2018, 2016; Kshirsagar et al., 2023; Liu et al., 2022; Tang et al., 2022). To evaluate the machine learning model’s accuracy, we examined the analyzer’s optical detection using synthetic dyes and real-time LAMP assays. We prepared six concentrations of FAM and HEX dye (0, 0.2, 0.4, 0.6, 0.8, 1 µM) across 36 potential combinations and collected ten data points per combination, replicated four times, and split into 80/20% for training and testing. We found our model performed very well like before(Kshirsagar et al., 2024b; Liu et al., 2025a), demonstrating very strong linearity for both FAM (R^2^ = 0.994) and HEX (R^2^ = 0.996) across all combinations of manual dye concentration (**Figure *3*b**). These results suggest our analyzer’s optical module and machine learning algorithm are well suited for the multiplex analysis of real-time LAMP assays. To verify the analyzer’s real-time LOD, we conducted several amplification assays on the device. In addition, we duplicated the assays to be operated in parallel on the benchtop thermal cycler. While RT-PCR and RT-LAMP have different amplification mechanisms, RT-PCR remains the gold standard in NAT(M. A. Ahamed et al., 2025). In **Figure *3*c**, we found that the portable analyzer showed great agreement with the benchtop device for both HIV (r = 0.985) and RNase P (r = 0.962) detection across a range of concentrations. Strong linearity and more than 96% agreement with the benchtop analyzer suggests the successful implementation of the previous NN model(Kshirsagar et al., 2024a) on our HIV analyzer is highly accurate, reproducible, and robust. These features make our device well suited for portable analysis of multiplex assays. Further validation of our device is needed to examine its performance detecting clinical plasma samples.

**Figure 3.**
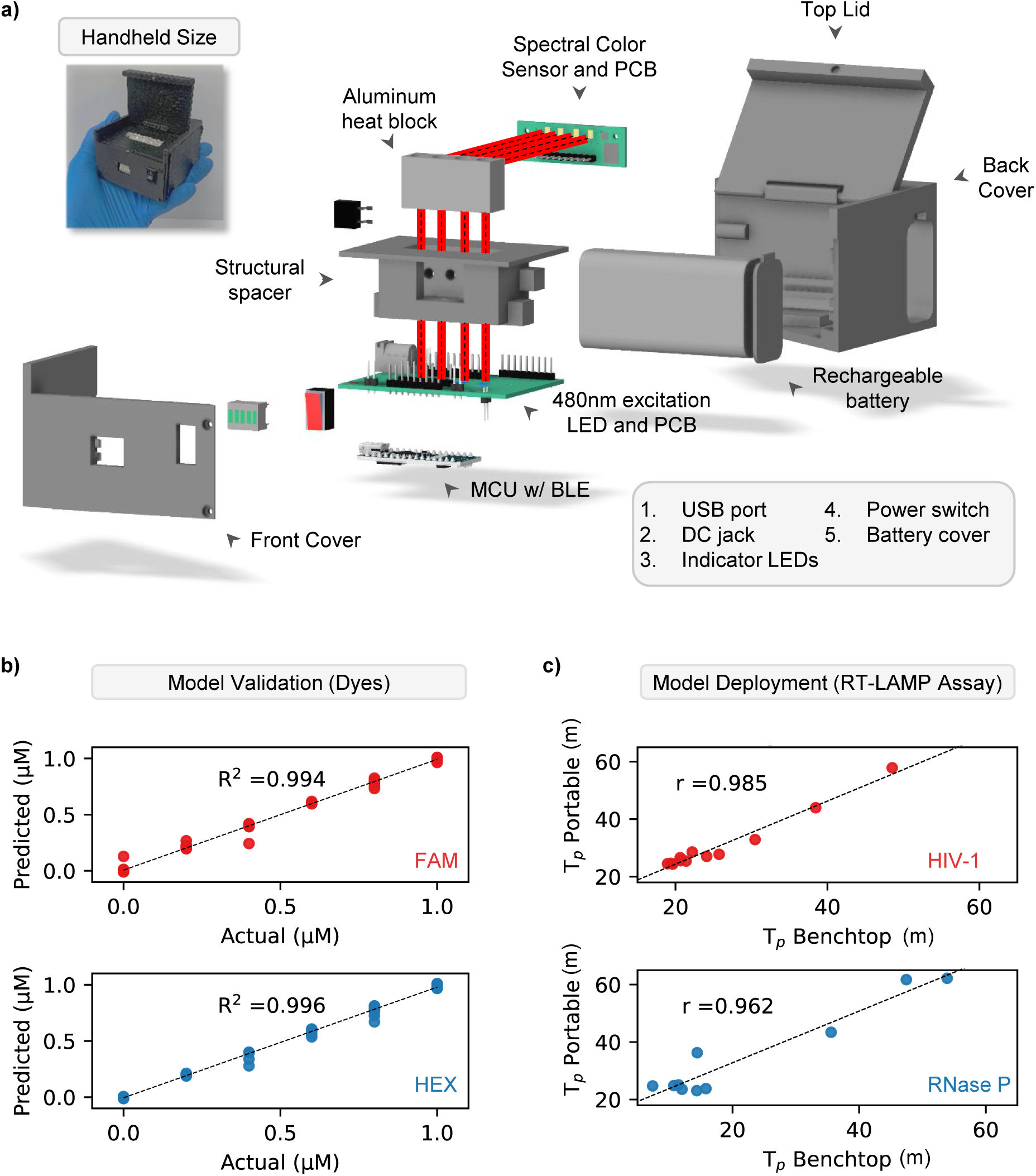
Validation of hardware and software of the ViraLite analyzer. a) The ViraLite analyzer is composed of several electrical components housed within a custom, 3D printed enclosure. The 480nm excitation LEDs are located on the horizontal mainboard located underneath the structural spacer and heat block. Light from these LEDs will travel vertically upward into the heat-block where PCR tubes are located with the assay. Once excited, the assay will emit photons, those are detected using the spectral color sensor located orthogonally to the path of excitation light (light path is highlighted in red). A BLE enabled MCU is located on the bottom of the mainboard PCB and connects the analyzer’s components together, monitors internal feedback, and transmits data. b) The analyzer and machine learning software (neural network) showed excellent performance detecting increasing dye concentrations (FAM and HEX). c) Last, the analyzer showed very strong agreement with the benchtop thermal cycler for analyzing both targets (HIV-1 and Rnase P) in multiplex format over a range of concentrations (10^5^ down to 10 copies).

### Deploying ViraLite for Qualitative testing

To validate our analyzer’s qualitative performance, we examined its clinical sensitivity and specificity using 45 plasma samples. Each sample was divided into two equal aliquots of 50 µl. One sample was prepared and analyzed using typical laboratory protocol (centrifuge & qRT-PCR) (**Figure *4*a**). The second aliquot was prepared and analyzed using our portable extraction methods and analyzed using our multiplex RT-LAMP analyzer (**Figure *4*b**). These results are summarized in **Figure *4*c** to provide qualitative analysis (data shown in **Supplementary Table S1**). Our PCR results indicated 21 positive and 24 negative samples respectively. Initially, our RT-LAMP results identified 15 positive and 30 negative samples, demonstrating a relatively poor sensitivity (66.66%) but high specificity (100%). However, the advantage of our internal quality control-based LAMP assay enables us to remove invalid tests, samples where no RNase P is detected. Regardless of HIV detection, a sample will be deemed invalid if no RNase P is detected. Removing these samples reduced our data set to 15 positive and 13 negative samples. As a result, our RT-LAMP performance greatly improves from 66.66% sensitivity to 93.33% as we identify 14 positive and 13 negative samples (**Figure *4*d**). The specificity of our assay remained 100%. With a sensitivity of 93.33% and 100% specificity, we demonstrate our analyzer’s strong qualitative ability for classifying clinical samples.

**Figure 4.**
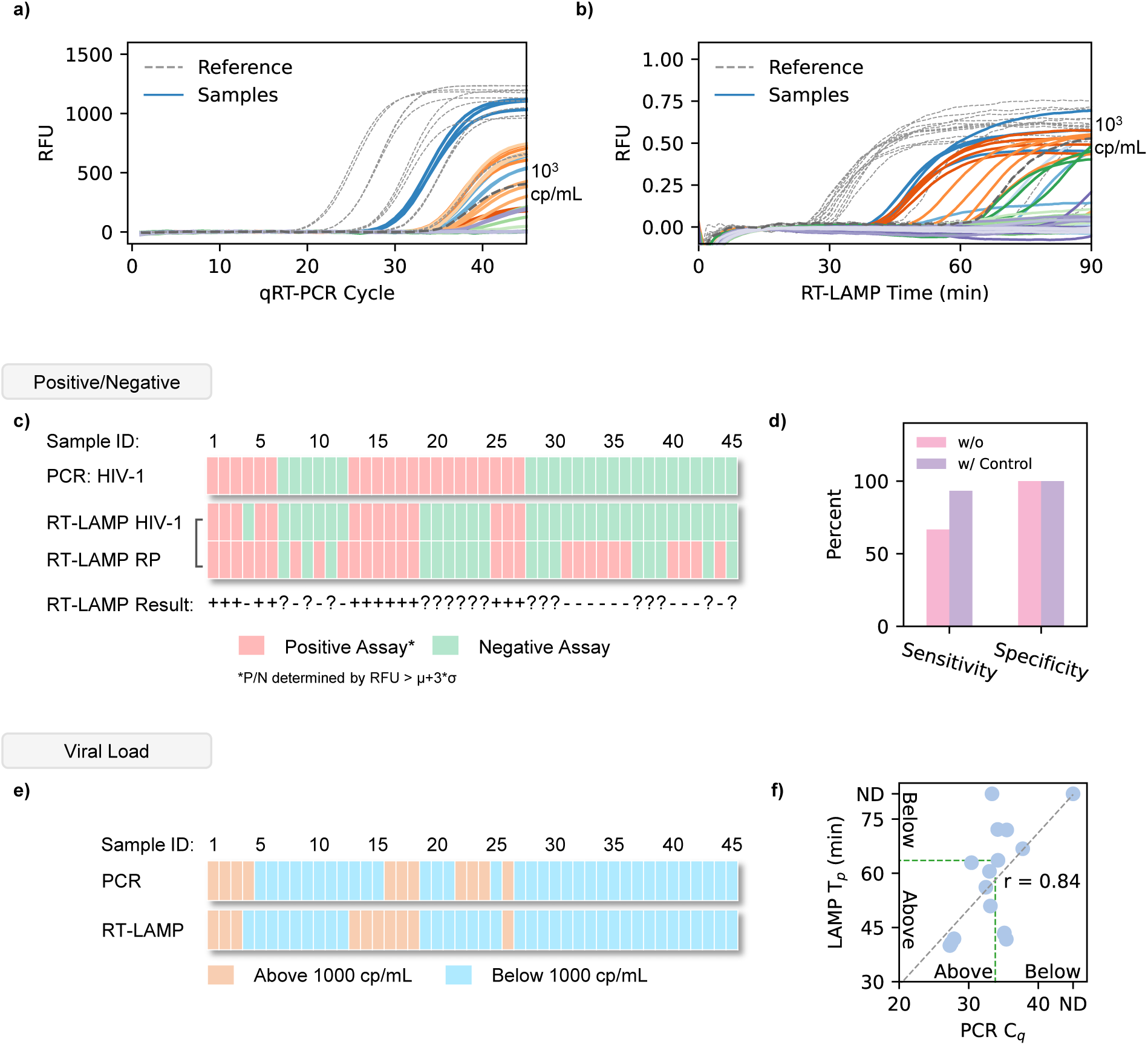
Clinical demonstration of ViraLite using Sensitivity, Specificity, and Semi-quantitative viral loads. a) Amplification curves from 45 clinical samples using qRT-PCR as a benchmark. A 1000 cp/mL marker is designated as a dark grey dashed line. b) Amplification curves from 45 clinical samples, analyzed using our multiplex LAMP assay on our portable analyzer (data smoothing applied with python). A 1000 cp/mL marker is designated as a dark grey line. c) Positive vs Negative comparison of qRT-PCR vs our multiplex LAMP output, displayed for all 45 samples. Positive assays are labeled in red. Negative assays are labeled in green. Positive/Negative was established using a RFU threshold of µ+3σ. d) Clinical sensitivity and specificity of our multiplex LAMP assay with and without an internal quality control to eliminate invalid samples. e) Viral load classification of qRT-PCR vs our multiplex LAMP assay for 45 clinical samples. The typical clinical threshold of 1000 cp/mL was used to define Above (>1000 cp/mL) and Below (<1000 cp/mL) classifications. f) LAMP T_p_ vs PCR C_q_ for all valid samples, demonstrating a strong correlation in analysis between both methods.

### Deploying ViraLite for Semi-Quantitative testing

To demonstrate ViraLite’s usability for semi-quantitative viral load monitoring, we classified each sample using qRT-PCR and our RT-LAMP. To achieve this, we classified each sample’s qRT-PCR C_q_ and RT-LAMP T_p_ based off the clinically recommended threshold for decreased transmission risk, 1000 cp/mL (Calmy et al., 2007; Eisinger et al., 2019; Farzadegan et al., 1998; Phillips et al., 2001; Saag et al., 1996; Wilson et al., 2008), This was established using five reference concentrations for both PCR and RT-LAMP (dashed lines in **Figure *4*a-b**) to create two classes: Above and Below. The classification is summarized in **Figure *4*e**. These reference concentrations showed much smaller standard deviations compared to actual samples because they were tested using purified HIV RNA versus extracted RNA from actual samples. Our analyzer identified 10 ‘above’ and 18 ‘below’ samples, whereas qRT-PCR classified 8 ‘above’ and 20 ‘below’. Comparing the analyzer’s T_p_ against the C_q_ from benchtop PCR (**Figure *4f***), we found our analyzer accuracy to be 86% and our overall quantitative ability showed 84% agreement with the benchtop device. These results are expected since our multiplex RT-LAMP assay shows an order of magnitude higher LOD than the typical qRT-PCR assay (1-10 cp/rxn). We expect that further improvements to this assay could boost its analytical sensitivity and therefore improve the overall accuracy for quantification. Currently, our analyzer boasts 93.33% sensitivity and 100% specificity, showcasing the capability and need for internal quality-controlled RT-LAMP assays. With an accuracy of 86% and 84% agreement with benchtop qRT-PCR, ViraLite demonstrates a capability for semi-quantitative viral load monitoring.

### Surveying ViraLite perception among patients in HIV clinics

To evaluate the potential acceptance and usability of our system, we surveyed 480 patients on HIV or PrEP treatment. Voluntary patients at several HIV clinics were asked to watch a short video (**Supplementary Video S2**) showing the steps to operate ViraLite. After watching the video, patients were asked a series of demographic and response questions. In **Figure *5*a-f** we summarized the patient demographics using gender, age, income, education level, urban/rural, and race. These demographics align with the 2023 U.S. Statistics for HIV, suggesting a widespread and unbiased survey (**Supplementary Information**) (“HIV & AIDS Trends and U.S. Statistics Overview,” n.d.). Next, survey responses were categorized by the treatment plan, HIV antiretroviral therapy or pre-exposure prophylaxis (PrEP) (**Figure *5*g**). We found that 55.6% of patients were on HIV treatment and the remaining 44.4% of patients were on PrEP. More than half of participants receiving HIV treatment indicated Convenience, Privacy, Confidentiality, and Time Saved as their most perceived benefits (**Figure *5*h**). Participants on PrEP were more interested in benefits associated with Time Saved, [decreased risk of] Transmission, and Privacy. Less than 50% of all participants were interested in the benefits of Time to results and Cost. On the other hand, survey participants on HIV treatment were most concerned with Accuracy, Guidance, and Prescription (**Figure *5*i**). Patients receiving PrEP showed highest concern with Prescription [management], Misinterpretation [of results], Accuracy, and Guidance. Less than half of all participants were concerned with Emotional Impact [of tests/results], Record Keeping, or Cost. These results suggest that participants are deeply interested in tests that bridge the gap between clinic care and at home testing; however, they are concerned about the accuracy and guidance of ViraLite. Notably, the ‘Cost’ of the device was neither a major benefit nor concern for people, suggesting the usefulness of at home monitoring outweighs the potential costs of ownership. Concerns over prescription management may suggest that patients are unsure how at-home testing will affect their prescription. This is an interesting topic area we will further develop alongside ViraLite. Overall, ViraLite agreement with survey feedback suggests there is high potential for immediate positive impact on patients. At the same time, the future development of ViraLite will heavily consider concerned areas as we attempt to translate this technology into patients’ hands.

**Figure 5.**
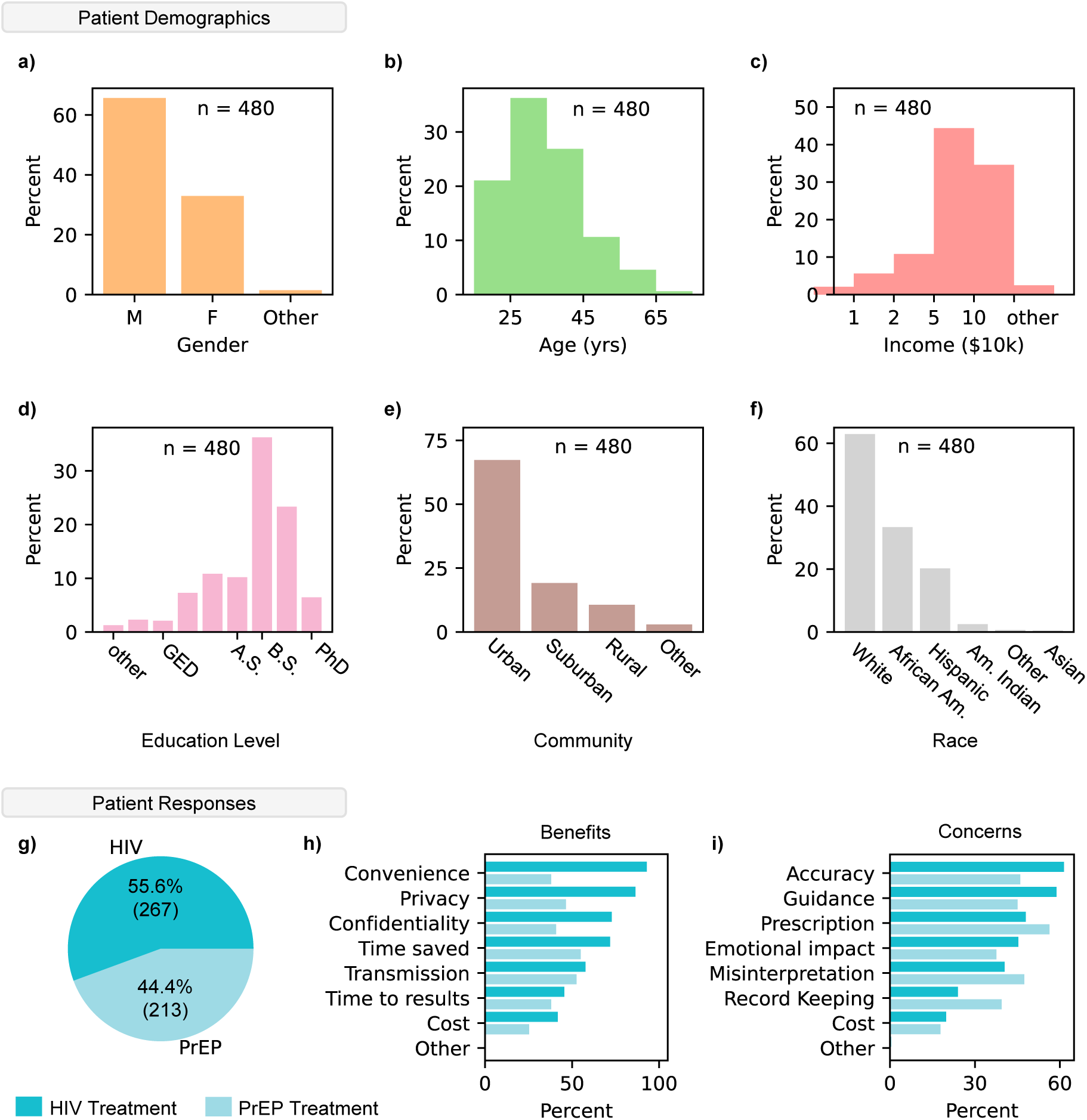
Evaluation of survey participant feedback toward ViraLite improvements. The percentage of participants (n = 480) for each demographic: a) Gender, b) Age, c) Income, d) Highest Education level, e) Community, and f) Race. g) Patient responses indicated a 55.6% (HIV) to 44.4% (PrEP) separation between current treatments. h) Percent indication of the perceived benefits of our system from patients on either treatment (HIV – dark, PrEP – light). The highest percentage of marked benefits were Convenience, Privacy, and Confidentiality (HIV treatment). i) Percent indication of the potential concerns of our system from patients on either treatment (HIV – dark, PrEP – light). The highest percentage of marked concerns were Accuracy, Guidance, and Prescription [management] (HIV treatment).

## Discussion

The number of PLHIV is expected to rise in the coming decade due to advancements in ART, increasing the need for effective VL management(J Buzón et al., 2010; Johnson et al., 2017; Peter et al., 2017). Newly developed HIV VL monitoring tests should ideally demonstrate excellent clinical sensitivity and specificity to protect both patient and public health(Drain et al., 2019b; Hayes et al., 2019). This is best achieved through timely & convenient testing to encourage patients and empower self-management(Mariani et al., 2020). However, large sample volumes and lab infrastructure remain key challenges for accessible VL monitoring(Dorward et al., 2018). To address these issues, there is a need for portable, sensitive, and specific assays to bring convenient VL monitoring closer to patients.ViraLite’s probe-based RT-LAMP assay provides a highly sensitive and specific system. With The integration of HIV VL monitoring, machine learning, and battery-powered analzyers bridges the gap between clinic and patient. (Kshirsagar et al., 2024a, 2023; Liu et al., 2023, 2022; Tang et al., 2022)On the other hand, sample preparation continues to be a challenge for compact devices, while lyophilization remains difficult for customized assays. To summarize, the ViraLite system presented here enables highly sensitive, portable, self-testing that is empowered by multiplex RT-LAMP, machine learning, and semi-automated, battery-powered electronics. ViraLite demonstrates the seamless integration of sample preparation, multiplex LAMP detection, and smartphone guidance for an accessible testing system for HIV VL monitoring. Probe-based RT-LAMP enables multiplexed assays to rival PCR assays in detection and monitoring clinical samples. Machine learning empowers ViraLite to use simple, off-the-shelf optical components for accurate quantification of one-pot multiplex assays. Clinical validation with clinically archived plasma samples demonstrated the real-world application of ViraLite’s by coupling reliable clinical data with easy to access, portable hardware. Survey participants reassured us of our target demographic and gave us defined areas of improvement. In summary, ViraLite introduces a viable monitoring system to translate VL management outside of clinics and into accessible, comfortable locations. The conveniency of our system has the potential to rapidly change the frequency of VL monitoring, offering improved ART, decreased transmission, and advanced drug development.

## Materials and Methods

### Multiplex RT-LAMP assay

The RT-LAMP assay reaction, optimized for the detection of HIV-1, involves a reaction mix with a total volume of 25 µl. The assay (shown in Supplementary Table S2) contains an isothermal buffer, which includes Tris-HCl, (NH_4_)_2_SO_4_, KCl, MgSO_4_, Tween 20, and dNTPs, along with *Bst*, PCR grade H_2_O and MgSO_4_ (6 mM) were also included in the reaction mixture. RNA template was added in addition to primers, consisting of 0.2 µM of F3 and B3, 1.6 µM of FIP and BIP, and 0.8 µM of LPF and LPB. The reaction was performed at 60 ℃ for a duration of 60 minutes. For the multiplexed assay, the primer concentrations were reduced to a ratio of 3:2 HIV to RNase P. The HIV primer concentrations were 0.15 µM F3 and B3, 1.2 µM FIP and BIP, 0.4 µM Fd and QFIP, and 0.6 µM LF and LB. The RNase P primer concentrations are 0.1 µM F3 and B3, 0.8 µM FIP and BIP, 0.4 µM LFP, Q and LB. All RT-LAMP primers and probes are listed in Supplementary Table S3.

### qRT-PCR Assay

The PCR HIV assay was previously validated by Palmer et al. (Palmer et al., 2003) (See Supplementary Table S4). The total PCR volume consisted of 25 μL: 6.25 μL of Fast Taq One-Step Master Mix (Applied Biosystems, Waltham, MA), 1.5 μL of Forward and Reverse primer, 0.63 μL of Probe, 10 μL of extracted RNA sample, and 5.13 μL of Nuclease-free water (New England Biolabs, Ipswich, MA). Analysis was conducted using a Bio-Rad C1000 Thermal Cycler (Hercules, CA). Thermal Cycling was set as 50 °C for 5 min, 95 °C for 3 s, 65 °C for 30s, and repeated 60×. Primers and probes were purchased from Integrated DNA Technologies (Coralville, IA). Positive samples were identified and tagged with a quantitative cycle (C_q_) value when the background RFU reached a threshold defined as: μ + 3σ.

### ViraLite instrumentation

The four sensor analyzer was designed using PTC Creo software and 3D-printed using a Method X printer from MakerBot system. The printed circuit boards (PCB) were designed using Autodesk Eagle and manufactured by OSH Park. Laboratory personnel soldered individual electronic components and the Microcontroller Unit (MCU) onto the PCB. The thermal module consists of a resistive-heating element (PWR263S-20-2R00J, Digi-Key) attached to the underside of the aluminum heating block using thermal compound (AATA-5G, Artic Alumina). Temperature feedback is provided from within the core of the heating block by a thermistor (95C0606, Digi-Key). Sustained temperature for extended periods of time was achieved using negative feedback control through an N-channel power Metal-Oxide-Semiconductor Field-Effect Transistor (MOSFET) (63J7707, Digi-Key). The optical module was designed using an adjustable Res Cermet Trimmer (3296W, Digi-Key) for LED intensity control, a wide spectrum IC Color Sensor (AS7341, Digi-Key) for optical detection, and a blue SMD LED (1497-1138-1, Digi-Key) for fluorescence excitation. An I^2^C multiplexer was used as the expander for the sensor. The smartphone graphical user interface (GUI) software was developed using MIT App Inventor. The GUI facilitates control and communication with the device via Bluetooth (**Supplementary Figure S6a-b**). A Bill of Materials for the analyzer is included as **Supplementary Table S5**.

### Sample preparation with portable device

Portable extractions were achieved using the Viral RNA Mini kit from Qiagen. Following our previously established protocol(Politza et al., 2024), extractions were performed with 50-100 µl sample, 500 µl AVL Buffer, 500 µl 95% Ethanol, 500 µl Wash Buffer I, 500 µl Wash Buffer II, 500 µl 95% Ethanol, and 80 µl water. During all extraction stages the portable device runs at max speed (6000 rpm/ 1743 rcf.). Our smartphone app will guide a user through the process of this extraction (Seen in Supplementary Video S1).

### Sample preparation with benchtop device

Benchtop extractions were conducted according to the Viral RNA Mini kit from Qiagen. Extractions were performed with 100-140 µl sample, 500 µl AVL Buffer, 500 µl 95% Ethanol, 500 µl Wash Buffer I, 500 µl Wash Buffer II, 80 µl water(Politza et al., 2024). The benchtop centrifuge was set to 1 min. at 6000 rcf. for all steps except Wash Buffer II where the centrifuge was operated for 3 min. at 20,000 rcf.

### Clinical Samples

Plasma samples were collected from HIV patients treated at the Penn State Hershey Medical Center following controlled standard operating procedure. Investigational Review Board (IRB) approval was provided by the Pennsylvania State University (STUDY00015905). The samples were de-identified and randomly selected from remnant plasma batches. No identifiable data was recorded or transferred between study locations. For each test, 50 µl of clinical plasma sample was used.

### Survey recruitment

Survey participants were recruited online and at the Penn State Hershey Medical Center, the UPMC infectious disease center, the Penn Health Lancaster General Health Comprehensive Care Center, Seattle King County HIV/STI clinics (Seattle, Washington), and the Washington University HIV/STI clinics (St. Louis, Missouri). In total 1200 surveys were collected from all site locations. We discarded any survey which did not watch more than 30s of the video, therefore excluding 720 respondents, leaving 480 respondents. Institutional Review Board (IRB) approval was provided by the Pennsylvania State University (STUDY00024241). Participants were compensated with a $10 Amazon gift card for the successful completion of the survey.

### Statistical analysis

All data processing and figure generation were completed using Python. Data is displayed as the mean of triplicates plus or minus three standard deviations, unless otherwise noted. Positive samples are classified using Time to Positive (T_p_) or quantitative cycle (C_q_) when RFU reaches a threshold of μ + 3σ. LOD was calculated with 95% confidence using a logistic function fit to the hit rate for each assay. Correlation and linearity were computed using SciPy and least squares regression.

## Data Availability

All data produced in the present study are available upon reasonable request to the authors.

## Acknowledgments

This work was partially supported by the National Institutes of Health (R61AI147419, R33AI147419), and the National Science Foundation (1912410, 2319913). Any opinions, findings, conclusions, or recommendations expressed in this work are those of the authors and should not be construed to represent any official NIH and NSF or U.S. Government determination or policy.

## Declarations

The authors declare the following competing financial interest(s): A provisional patent has been filed related to the technology described herein.

## Author Contributions

**Anthony J. Politza**: conceptualization, software, investigation, writing – original draft. **Tianyi Liu**: conceptualization, resources, writing – review & editing. **Aneesh Kshirsagar**: conceptualization, resources, writing – review & editing. **Ming Dong**: methodology, supervision, visualization, writing – review & editing. **Md. Ahasan Ahamed**: visualization, writing – review & editing. **Muhammad Asad Ullah Khalid**: supervision, writing – review & editing, project management. **Roland Jones**: resources, methodology, and investigation. **UttaraSeshu**: formal analysis, data curation, writing – review & editing. **Kathryn Risher**: formal analysis, data curation, writing – review & editing. **Casey N. Pinto**: data curation, project administration, supervision, writing – review & editing. **Yusheng Zhu**: investigation, methodology, resources, supervision. **Weihua Guan**: conceptualization, supervision, writing – review & editing, project administration, and funding acquisition.

## Notes

### Funding Statement

This study was partially funded by the National Institutes of Health (R61AI147419, R33AI147419), and the National Science Foundation (1912410, 2319913). Any opinions, findings, conclusions, or recommendations expressed in this work are those of the authors and should not be construed to represent any official NIH and NSF or U.S. Government determination or policy.

### Author Declarations

IRB of Pennsylvania State University gave ethical approval for this work (STUDY00015905) IRB of Pennsylvania State University College of Medicine gave ethical approval for this work (STUDY00024241)

### Summary of Updates

In the first section, we revised the Author names and included some essential references in the introduction section.

