## Supplementary Information for "ViraLite: An Ultracompact HIV Viral Load Self-Testing System with Internal Quality Control" (from another version of this preprint: version 1, 2025-04-03)

**RT-LAMP Design.** Earlier studies into multiplexed LAMP have demonstrated probe-based detection using labeled primers[1–3]. Detection of Amplification by Releasing of Quenching (DARQ) supplements the standard LAMP primers with a duplex probe[4,5]. In this method the duplex consists of a quencher attached forward inner primer (QFIP) annealed to a fluorophore labeled F1 region (Fd)[4]. Another method of probe-based LAMP, 5p-OSD, also supplements the standard LAMP assay with a duplex probe[6,7]. The difference between DARQ and 5p-OSD is apparent in the utilization of one of the standard loop primers (LP) as one half of the added duplex[7]. In this manner, 5p-OSD is designed with a 5’-fluorophore and annealed to a complementary strand containing a 3’ quencher (Qd) . Our design uses both DARQ (HIV-1) and 5p-OSD (RNase P) to achieve a multiplex LAMP assay for HIV-1 and RNase-P.

**Analyzer Design.** In a previous study we showed that using single color channels to monitor bulk assay fluorescence limited the capabilities of multiplexing [8] (**Supplementary Figure S5**). However, using a multi-spectra sensor enables ViraLite to analyze broad spectrum output from an assay and then a trained machine learning model can assist in the conversion from 8 channel signal to 2 fluorophore concentrations. The handheld device is constructed using 3D-printed ABS to support the electrical and thermal modules. Inside the device, a 7.4V 2200mAh Li-ion battery provides power to the motherboard, resistive heater, and Arduino MCU. The MCU controls the excitation LEDs, indication LEDs, resistive heating module, and optical array. Excitation light from the blue LEDs (480 nm) enters the heating block from the bottom where the incident light hits the assay tube. Then, emitted light from the assay is detected by color sensors positioned 90° off axis of the excitation light.

**Survey information.** A significant majority of our survey participants were male (60%) with a minority population of female and other (transgender male, transgender female, non-binary, other). We found most of our participants were between 25-34 with an average income between $50,000-99,000. Our results showed that for the plurality of participants the highest education level among participants was a bachelor’s degree. The most common environment for respondents was an urban setting and our surveyed population was primarily white with small but present representation of African American and Hispanic populations.

### Supplementary Figures

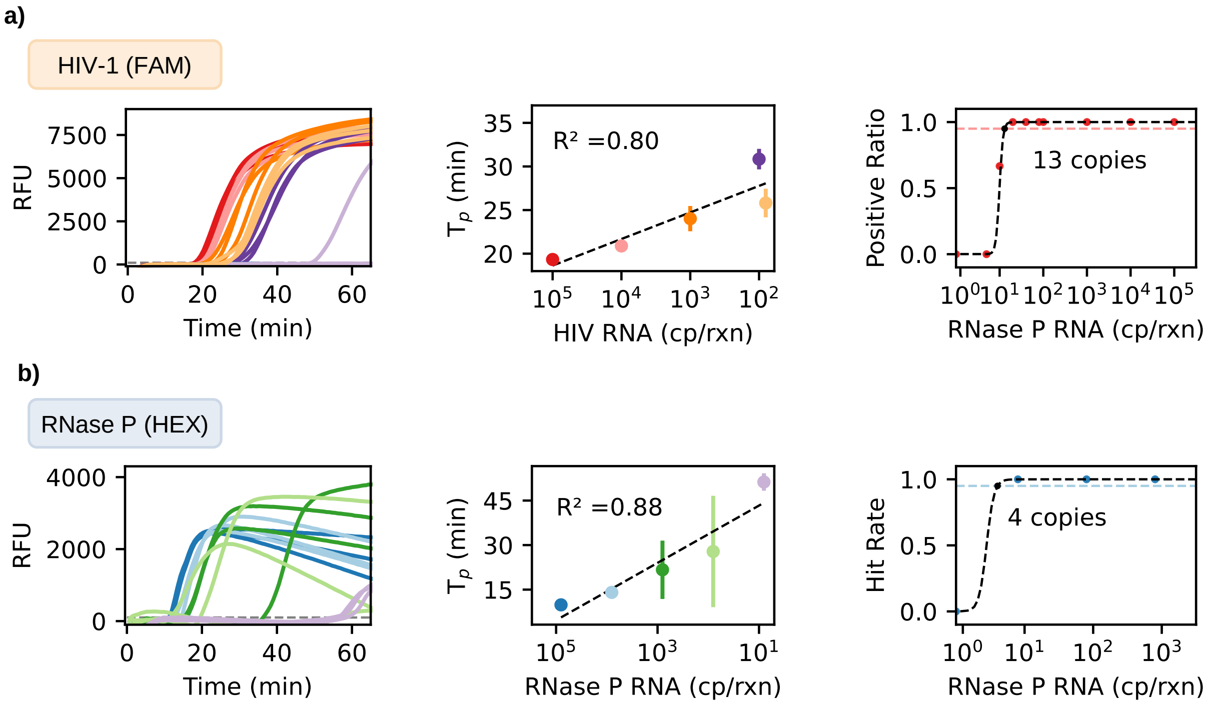

**Figure S1.** Single-plex assay validation. a) The probe-based HIV-1 Assay was initially screened using $10^5$ down to 1 copy of viral RNA. The summary of detection linearity showed an $R^2$ of 0.80 and resulted in a limit of detection of 13 copies (95% CI). b) Similarly, the probe-based RNase P assay was screened using $10^5$ down to 1 copy of viral RNA. The summary of detection linearity showed an $R^2$ of 0.88 and resulted in a limit of detection of 4 copies (95% CI).

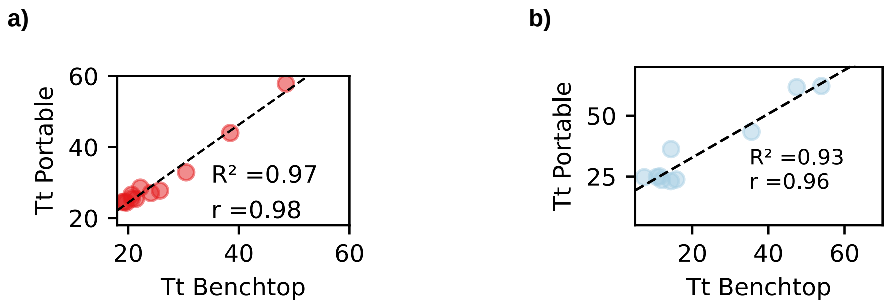

**Figure S2.** Single-plex correlation using portable vs benchtop. a) Our HIV-1 assay showed very strong correlation (r = 0.98) between analyzer formats and great linearity ($R^2$ = 0.97) for semi-quantitative analysis. b) Our RNase P assay also showed very strong correlation (r = 0.96) between analyzer formats and great linearity ($R^2$ = 0.93) showcasing the potential for multiplex detection between both assays, regardless of sensitivity losses.

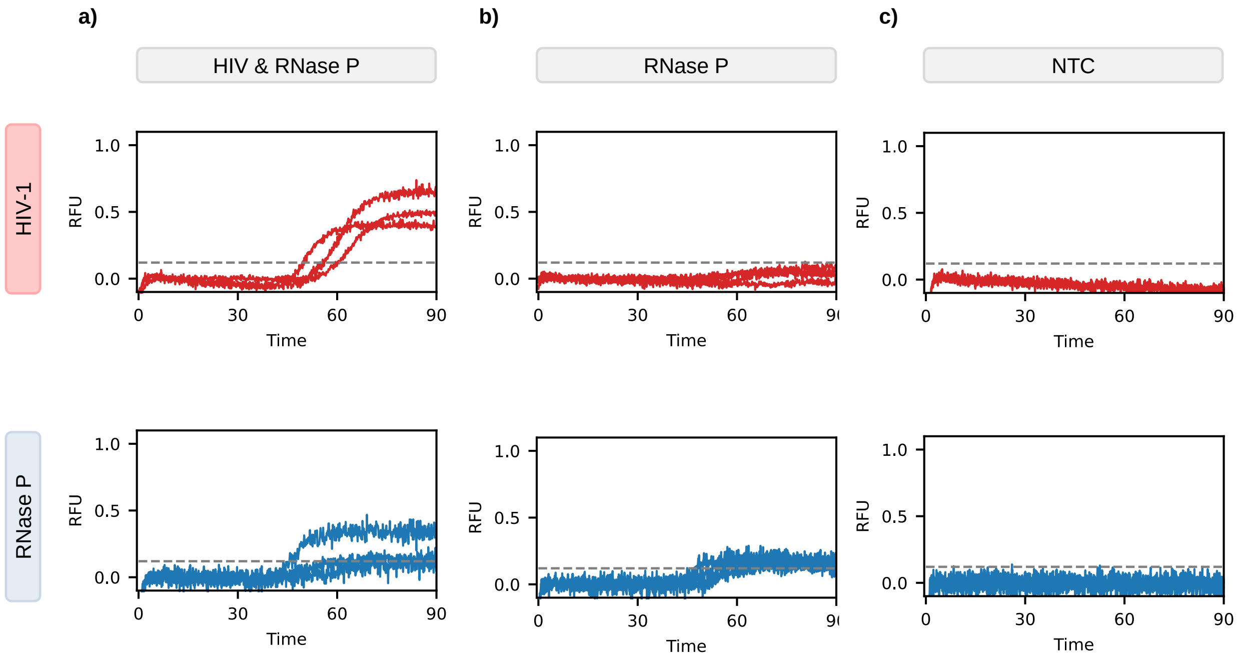

**Figure S3.** Multiplex cross-reactivity using our portable analyzer. a) The multiplex assay used FAM and HEX fluorophores to distinguish HIV-1 and RNase P, respectively. When both targets are present in the sample, there is exponential amplification for both FAM and HEX. b) For samples with only RNase P, there is only amplification detected in the HEX channel. c) No Target Controls (NTC) show no amplification for either FAM or HEX (HIV-1 or RNase P).

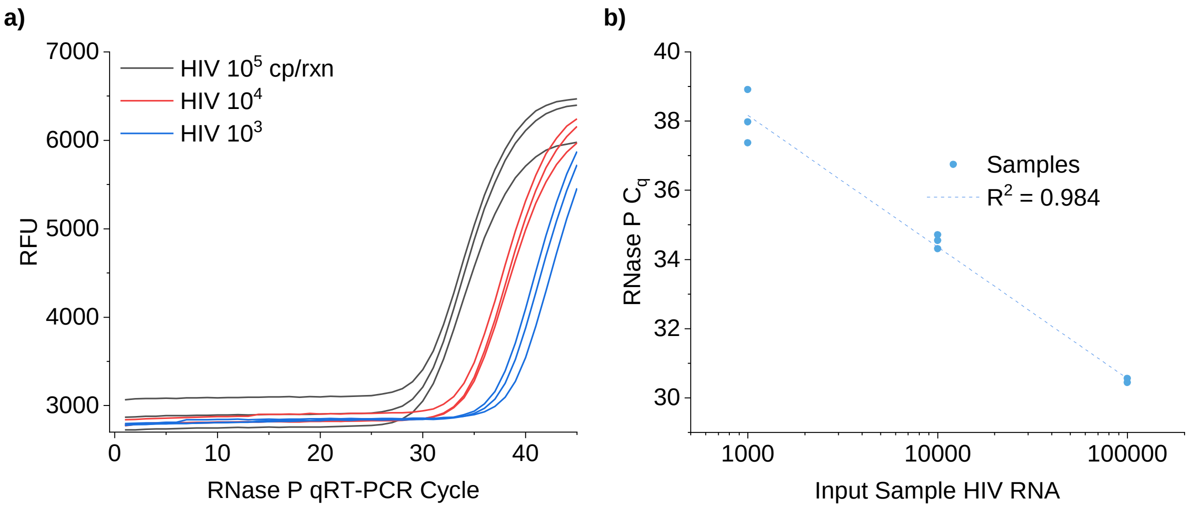

**Figure S4.** RNase P Validation. a) qRT-PCR amplification curves from HIV Seracare samples at three diluted concentrations, showing RNase P presence. b) PCR Cq showed a linear relationship against input concentration.

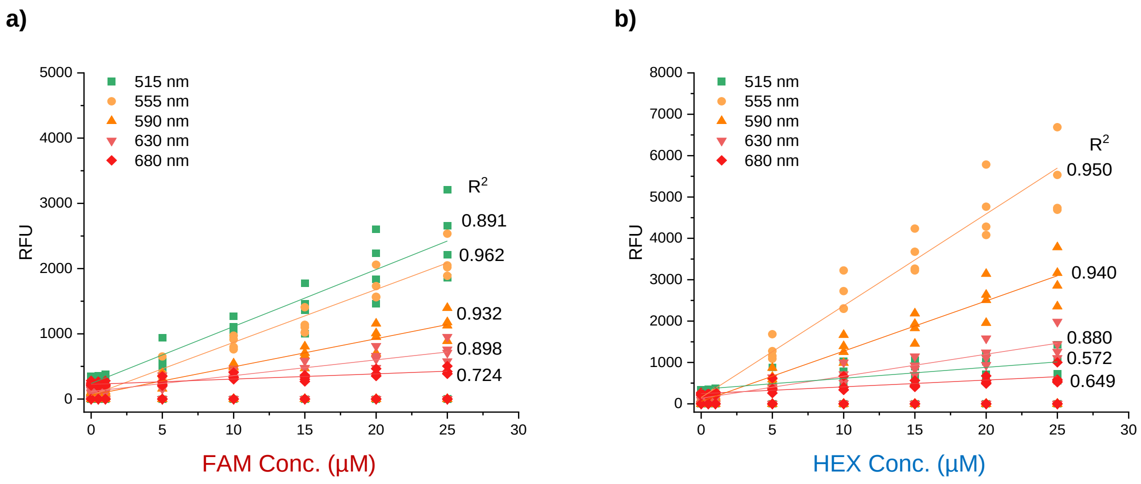

**Figure S5.** Typical dye performance using single channel detection. a) Color sensor readout from five channels showing the crowded yet accurate detection of increasing FAM dye. b) Identical five channel output but now for increasing concentrations of HEX dye. When used in combination the two dyes are indistinguishable, given that both the 515 nm centered (FAM best) and the 555 nm centered (HEX best) channels are sensitive to the other dye.

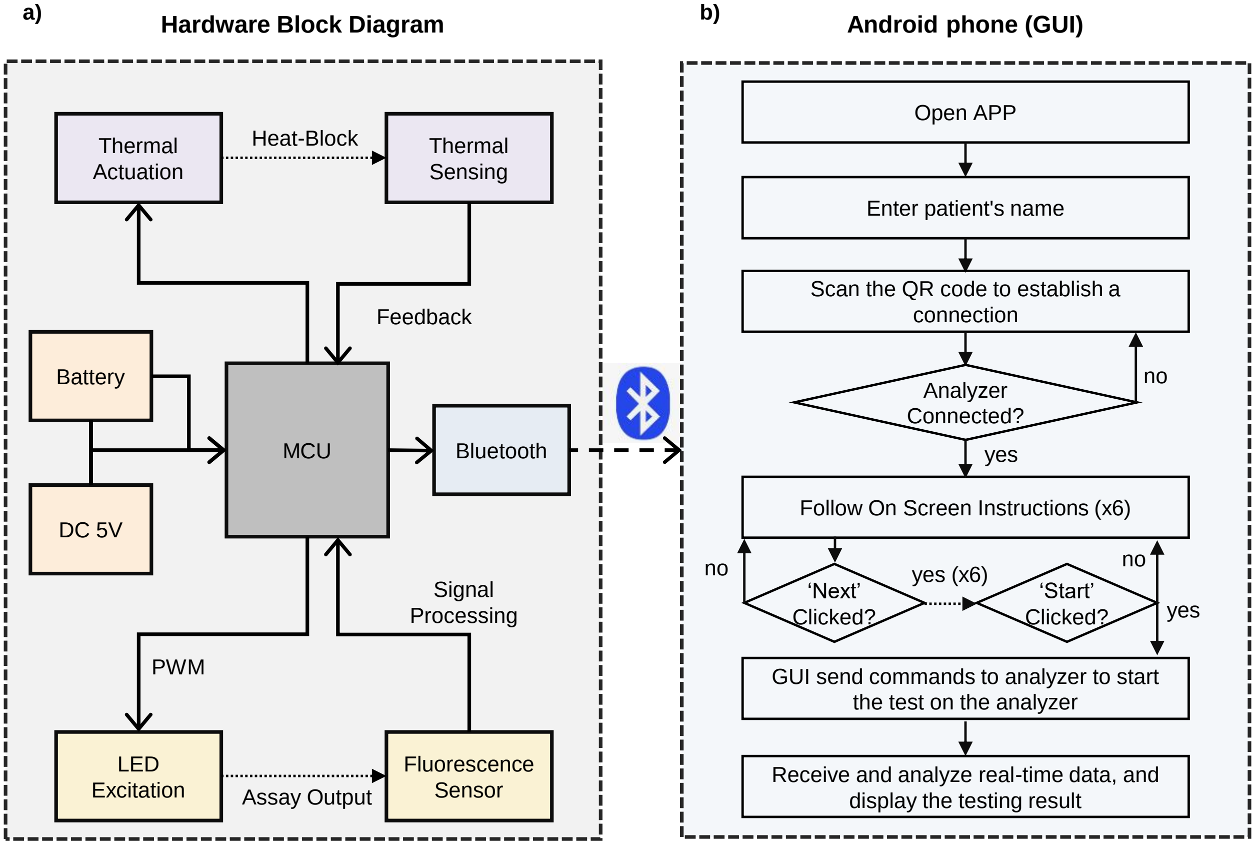

**Figure S6.** Software and smartphone app. a) Block diagram illustrating the interconnecting components of the portable analyzer. Output includes a resistive heater, an LED for excitation, and a status LED bar. Inputs include temperature feedback from the thermistor and an a wide-spectra color sensor. An Arduino Nano controls all modules and interfaces with an Android phone through Bluetooth. b) Block diagram demonstrating the workflow of the Android smartphone app. The GUI will walk patients through the entire testing workflow, confirming each step has been completed, and will display results on the last page with an option to upload to cloud storage sites.

### Supplementary Tables

#### Table S1. Clinically archived sample performance

| Sample Number | 1 | 2 | 3 | 4 | 5 | 6 | 7 | 8 | 9 | 10 | 11 | 12 | 13 | 14 | 15 | 16 | 17 | 18 | 19 | 20 | 21 | 22 |
|---|---|---|---|---|---|---|---|---|---|---|---|---|---|---|---|---|---|---|---|---|---|---|
| RT PCR HIV | 28 | 27 | 27 | 33 | 34 | 34 | 0 | 0 | 0 | 0 | 0 | 0 | 35 | 35 | 35 | 33 | 32 | 33 | 35 | 35 | 36 | 33 |
| RT PCR HIV | + | + | + | + | + | + | - | - | - | - | - | - | + | + | + | + | + | + | + | + | + | + |
| RT LAMP HIV | 42 | 41 | 40 | 0 | 72 | 64 | 0 | 0 | 0 | 0 | 0 | 0 | 43 | 42 | 43 | 51 | 56 | 61 | 0 | 0 | 0 | 0 |
| RT LAMP HIV | + | + | + | - | + | + | - | - | - | - | - | - | + | + | + | + | + | + | - | - | - | - |
| RT LAMP RP | 47 | 45 | 43 | 65 | 75 | 44 | 0 | 53 | 0 | 63 | 0 | 72 | 51 | 48 | 51 | 61 | 61 | 57 | 0 | 0 | 0 | 0 |
| RT LAMP RP | + | + | + | + | + | + | - | + | - | + | - | + | + | + | + | + | + | + | - | - | - | - |

| Sample Number | 23 | 24 | 25 | 26 | 27 | 28 | 29 | 30 | 31 | 32 | 33 | 34 | 35 | 36 | 37 | 38 | 39 | 40 | 41 | 42 | 43 | 44 | 45 |
|---|---|---|---|---|---|---|---|---|---|---|---|---|---|---|---|---|---|---|---|---|---|---|---|
| RT PCR HIV | 33 | 34 | 38 | 30 | 35 | 0 | 0 | 0 | 0 | 0 | 0 | 0 | 0 | 0 | 0 | 0 | 0 | 0 | 0 | 0 | 0 | 0 | 0 |
| RT PCR HIV | + | + | + | + | + | - | - | - | - | - | - | - | - | - | - | - | - | - | - | - | - | - | - |
| RT LAMP HIV | 0 | 0 | 63 | 67 | 72 | 0 | 0 | 0 | 0 | 0 | 0 | 0 | 0 | 0 | 0 | 0 | 0 | 0 | 0 | 0 | 0 | 0 | 0 |
| RT LAMP HIV | - | - | + | + | + | - | - | - | - | - | - | - | - | - | - | - | - | - | - | - | - | - | - |
| RT LAMP RP | 0 | 0 | 62 | 59 | 49 | 0 | 0 | 0 | 58 | 51 | 55 | 70 | 45 | 49 | 0 | 0 | 0 | 65 | 50 | 60 | 0 | 55 | 0 |
| RT LAMP RP | - | - | + | + | + | - | - | - | + | + | + | + | + | + | - | - | - | + | + | + | - | + | - |

#### Table S2. Multiplex LAMP assay

| Component | Volume (µL)/rxn | Final Concentration |
|---|---|---|
| NEB Isothermal Buffer | 5.00 | 1 X |
| HIV-F3 | 0.38 | 0.15 µM |
| HIV-B3 | 0.38 | 0.15 µM |
| HIV-FIP | 0.80 | 0.8 µM |
| HIV-BIP | 1.20 | 1.2 µM |
| HIV-LB | 0.60 | 0.6 µM |
| HIV-LF | 0.60 | 0.6 µM |
| HIV-QFIP:Fd | 0.40 | 0.4 µM |
| RNase P-F3 | 0.25 | 0.1 µM |
| RNase P-B3 | 0.25 | 0.1 µM |
| RNase P-FIP | 0.80 | 0.8 µM |
| RNase P-BIP | 0.80 | 0.8 µM |
| RNase P-LFP:Q | 0.40 | 0.4 µM |
| RNase P-LB | 0.40 | 0.4 µM |
| Betaine | 4.00 | 0.4 M |
| MgSO4 | 3.00 | 6 mM |
| dNTP Mix | 7.00 | 1.4 mM |
| Bst2.0 polymerase | 3.44 | 0.55 U/µl |
| Warmstart Reverse Transcriptase | 1.10 | 0.33 U/µl |
| RNA | 10.00 | 10 µl/rxn |
| UltraPure Water | 9.21 | - |

### Table S3. Probe-based RT-LAMP primers

| Target | Primer | Sequence (5' -> 3') | GenBank Target |
|---|---|---|---|
| RNase P | F3 | TTGATGAGCTGGAGCCA | U94316.1 |
| RNase P | B3 | CACCCTCAATGCAGAGTC | U94316.1 |
| RNase P | FIP | GTGTGACCCTGAAGACTCGGTTTTAGCCACTGACTCGGATC | U94316.1 |
| RNase P | BIP | CCTCCGTGATATGGCTCTTCGTTTTTTTCTTACATGGCTCTGGTC | U94316.1 |
| RNase P | LFP | /5HEX/ATGTGGATGGCTGAGTTGTT | U94316.1 |
| RNase P | LB | CATGCTGAGTACTGGACCTC | U94316.1 |
| RNase P | Q | CAGCCATCCACAT/3BHQ_1/ | U94316.1 |
| HIV-1 | F3 | AGTTCCCTTAGATAAAGACTT | AF033819.3 |
| HIV-1 | B3 | CCTACATACAAATCATCCATGT | AF033819.3 |
| HIV-1 | FIP | GTGGAAGCACATTGTACTGATATCTTTTTGGAAGTATACTGCATTTACCAT | AF033819.3 |
| HIV-1 | BIP | GGAAAGGATCACCAGCAATATTCCTCTGGATTTTGTTTTCTAAAAGGC | AF033819.3 |
| HIV-1 | LB | GCATGACAAAAATCTTAGA | AF033819.3 |
| HIV-1 | LF | GGTGTCTCATTGTTTATACTA | AF033819.3 |
| HIV-1 | QFIP | /5IABkFQ/GTGGAAGCACATTGTACTGATATCTTTTTGGAAGTATACTGCATTTACCAT | AF033819.3 |
| HIV-1 | Fd | GATATCAGTACAATGTGCTTCCAC /36-FAM/ | AF033819.3 |

### Table S4. qRT-PCR primers

| Target | Primer | Sequence (5' - 3') |
|---|---|---|
| HIV-1 pol | Forward | CATGTTTTCAGCATTATCAGAAGGA |
| HIV-1 pol | Reverse | TGCTTGATGTCCCCCCACT |
| HIV-1 pol | Probe | FAM-CCACCCCACAAGATTTAAACACCATGCTAA-BHQ1 |

### Table S5. Portable Analyzer Bill of Materials

| System | Description | Part# | Function | Unit Cost ($) | Unit Qty. | Ext. Cost ($) |
|---|---|---|---|---|---|---|
| Electronics | Arduino Nano | 7630049200173 | Microcontroller | 6.00 | 1 | 6.00 |
| Electronics | Adafruit Bluefruit LE SPI | 2633 | Bluetooth Low Energy | 14.00 | 1 | 14.00 |
| Electronics | Capacitor 10uF | P10425CT | Power stabilizing | 0.15 | 1 | 0.15 |
| Electronics | DC Barrel Power Jack/Connector | PRT-00119 | Power Connector | 1.25 | 1 | 1.25 |
| Electronics | 36-pin stripe male header | 392 | Headpins | 4.95 | 0.1 | 0.50 |
| Thermal | Aluminum CNC block | CP-0.91-0.91 | Heating Stage | 5.75 | 0.25 | 1.44 |
| Thermal | 2Ω Power Resistor | MP725-2.00-FCT | Heater | 7.01 | 1 | 7.01 |
| Thermal | SMD Resistor, 1kΩ | 311-1.00KHRCT | Temperature/EM control | 0.01 | 1 | 0.01 |
| Thermal | Thermistor | 235-1057 | Temperature sensing | 3.50 | 1 | 3.50 |
| Thermal | SMD Resistor, 10kΩ | RNCP0603FTD10K0CT | Temperature control | 0.10 | 1 | 0.10 |
| Thermal | N Channel Power MOSFET | DMN1019USN-7DICT | Switch for heater and fan | 0.35 | 2 | 0.70 |
| Optics | Res Cermet Trimmer 500K ohm | 3296W-1-501RLF | Adjust LED intensity | 0.60 | 3 | 1.80 |
| Optics | SMD Resistor, 1kΩ | RNCP0805FTD1K0CT | Optical module | 0.03 | 3 | 0.08 |
| Optics | SMD Resistor, 10kΩ | RNCP0805FTD10K0CT | Optical module | 0.03 | 8 | 0.22 |
| Optics | IC Color Sensor | AS7341 | Optical detection | 3.00 | 3 | 9.00 |
| Optics | SMD LED, Blue | 1497-1138-1 | Fluorescence excitation | 0.53 | 3 | 1.58 |
| Optics | Capacitor 10uF | P10425CT | Power stabilizing | 0.15 | 3 | 0.44 |
| Optics | Capacitor 0.1uF | P01425CT | Power stabilizing | 0.15 | 1 | 0.15 |
| Optics | Voltage Regulator | AP7312-3.3/1.8 | Power supplier | 0.80 | 1 | 0.80 |
| Optics | I2C Multiplexer | TCA9548A | I2C expender for sensor | 1.10 | 1 | 1.10 |
| Enclosure | ABS Filament (Black) | - | 3D platform material | 69.00 | 0.05 | 3.45 |
| Enclosure | Screws (M2 cap screw) | 91290A015 | Assembly | 0.14 | 10 | 1.40 |
| | | | | | **Total Cost** | **$63.54** |

### Supplementary Videos

**Supplementary Video S1. (Duration 2m 57s)** Demonstration of the testing workflow, using an updated version of RNA extraction.

**Supplementary Video S2. (Duration 5m 58s)** Initial demonstration of the testing workflow, shown to survey participants prior to the multiple select survey questions.
